# Childhood outcomes in children with Hirschsprung disease: a population-based data linkage study in England

**DOI:** 10.1101/2025.10.16.25338103

**Authors:** Katie L. Harron, Benjamin Jevans, Benjamin SR Allin, Joseph R. Davidson, Anestis Tsakiridis, Peter W. Andrews, Paolo De Coppi, Conor J. McCann

## Abstract

**Objective:** Hirschsprung disease (HSCR) is a rare congenital intestinal condition that, despite lifesaving surgery, can result in increased hospitalisations and a lower quality of life throughout childhood and into adulthood. There is a lack of population-level research on the reasons for these admissions, or on the level of support required at school age.

**Design/Methods:** We used linked administrative data from health and education services (ECHILD) to create a cohort of births from 2002-2020. We evaluated admission and mortality rates, number of surgical procedures, and reasons for admissions for children with and without HSCR at ages 0-4, 5-9 and 10-14. We assessed how many children had recorded Special Educational Needs by Year 1 of primary school (age 6).

**Results:** Of the 11,261,227 children in our cohort, 3227 (0.03%) had HSCR. 95.0% of children with HSCR were readmitted by age 4 compared with 40.2% of those without. Across ages and sexes, children with HSCR were frequently admitted for constipation, gastroenteritis, intestinal infections, abdominal pain, and nausea and vomiting. 3.2% of children with HSCR died by age 4 compared with 0.5% of children without HSCR. 44.0% of children with HSCR had recorded Special Educational Needs by age 6 compared with 17.9% of those without HSCR.

**Conclusion:** Children with HSCR have more frequent hospital admissions, surgical procedures, and higher mortality rates up to age 14 than their peers, and are more likely to have Special Educational Needs at school entry, irrespective of common comorbidities such as Down Syndrome. Improvements to treatment success, including novel or complementary approaches, are required in order to improve quality of life for this group of children. Further research is needed to understand the impact of HSCR and its treatment at the transition from paediatric to adult services, and on child development, progress in school, and other psychosocial factors including mental health.

**What is already known on this topic:** Hirschsprung disease (HSCR) is a rare congenital condition treated surgically in infancy, but long-term outcomes remain poorly understood. Existing studies have provided limited insight into broader population impacts of HSCR. Hence, comprehensive national data are needed to clarify ongoing health and developmental challenges in children with HSCR.

**What this study adds:** Using linked national health and education data from over 11 million children in England, this study provides the first population-level evidence on long-term outcomes in HSCR. Children with HSCR had significantly higher rates of hospital admissions, mortality, and Special Educational Needs compared with peers. These findings highlight the persistent health and developmental burdens of HSCR despite early surgical intervention.

**How this study might affect research, practice or policy:** Our study emphasises the need for coordinated long-term, multidisciplinary, care for children with HSCR. Our data can inform clinical pathways and policy planning to better address ongoing medical and educational needs for children and families living with HSCR.

## Introduction

Hirschsprung disease (HSCR) is a rare congenital intestinal condition that affects approximately 1 in 5000 live births.[1] The hallmark pathology is absence of ganglion cells within the enteric nervous system (the intrinsic innervation of the gastrointestinal tract) in a variable length of the distal bowel and occasionally even part of the small intestine.[2]

Most affected individuals are diagnosed in the neonatal period, though a proportion of patients may present in childhood with typical symptoms of severe constipation with abdominal distension and failure to thrive.[3] Treatment for the condition is surgical, involving the removal of the aganglionic bowel and performing a pull-through and anastomosis between ganglionic proximal bowel and the anorectum.[4, 5] This is achieved most commonly using either Duhamel or endorectal pull-through.[6]. Although primary surgery is generally the first choice among surgeons, there may be cases where a defunctioning stoma is necessary to safely decompress the bowel prior to elective surgery. Additionally, a proportion of affected individuals may have associated anomalies that mean faecal continence is likely to be poor and, in such cases, a definitive levelling stoma may be preferred.[7]

Although survival is high (>97%), children with HSCR are known to have higher morbidity than the general population, experiencing more frequent hospitalisations and longer inpatient stays, and increased requirement for surgery or procedures under general anaesthesia compared with children with other congenital anomalies or those without congenital anomalies.[8] In the longer-term, children with HSCR often experience lower quality of life than other children, and are commonly affected by ongoing bowel dysfunction, including faecal incontinence, chronic constipation, and soiling. One third of patients will require a further unplanned abdominal procedure.[9] While bowel function improves with age, >10% of children ultimately require a permanent stoma, the need for which has been identified as a core outcome of importance in determining the success of treatment for HSCR.[10–12] A third of patients report social problems related to bowel function into adulthood.[13]

The developmental outcomes of children with HSCR are less well studied. There is some evidence to suggest that children with gastro-intestinal anomalies, including HSCR, have impaired developmental and cognitive outcomes, which may be in part explained by frequent, extended hospitalization and numerous surgical procedures.[14, 15] There are concerns that multiple prolonged exposure to general anaesthesia during the neonatal period and infancy may affect brain development.[16, 17]

Although the epidemiology of HSCR has been well described in different clinical cohorts across Europe and North America, there is relatively little population-level research to understand the longer-term outcomes, including the need for educational support, of HSCR.[1] In this study, we used national administrative data from ECHILD (Education and Child Health Insights from Linked Data) to describe characteristics and outcomes of children with HSCR, in England, including hospitalisation outcomes up to 14 years of age and recording of Special Educational Needs by age 6.

## Methods

### Data source

We used ECHILD, which links administrative data from Hospital Episode Statistics (HES) and the National Pupil Database (NPD) for all children in England.[18] Education data from the NPD contains information on children attending state-funded schools in England, including Special Educational Needs (SEN).[19] Health data from HES analysed in this study included records of inpatient hospital attendances (Admitted Patient Care) at all NHS funded hospitals in England.[20] HES records contain basic demographic information (e.g., sex, ethnicity), area-level deprivation measured by the Index of Multiple Deprivation (IMD) of residential postcode, and clinical information based on International Classification of Diseases 10th Revision (ICD-10) diagnostic codes and Office of Population Censuses and Surveys Classification of Interventions and Procedures 4th Revision (OPCS-4) procedure codes. Information on birth characteristics such as gestational age, birthweight, and maternal age are also included through a mother-baby link.[21] Approximately 97% of all children born in NHS-funded hospitals in England have a birth record in HES. We also used linked mortality data from the Office for National Statistics (ONS).

### Study population

Our study included all singleton children born in NHS-funded hospitals between 1^st^ April 2002 and 31^st^ March 2020. Higher-order births (e.g. twins) were excluded due to less reliable linkage of hospital records over time. Hospital admission records from birth were used to identify a cohort of children with HSCR. For each admission, one primary diagnosis and up to 19 secondary diagnoses are recorded. Any recording of an ICD-10 diagnostic code for HSCR (Q43.1) was used to identify the cohort of children with the disease. This code has been shown to have 96% sensitivity and >99.99% specificity for identifying children with HSCR.[22] All other children (including those with other health conditions) were included in the comparison cohort.

Aligning with a previous study of HSCR using HES, we also identified the presence of common comorbidities based on ICD-10 codes: major cardiac anomalies (Q20-Q26) and chromosomal abnormalities (Q90-93, Q96-Q99, excluding Q93.6).[23]

### Outcomes

Our primary outcome of interest was admissions between ages 0-4, 5-9 and 10-14 years (excluding the birth episode). We also explored reasons for admissions, number of surgical procedures (identified through OPCS codes, excluding minor procedures, diagnostic imaging procedures, and codes describing subsidiary procedures/body parts, **Supp. Table 1**), and mortality. Focus groups and stakeholder work contributing to development of the Children’s Surgery Outcome Reporting (CSOR) programme have highlighted how the total number of operations a child undergoes is important to key stakeholder groups, not simply the number of operations that are related to a specific aspect of the child’s care or performed in a specific setting. Development of this programme has also highlighted how, without individual case review, it is not currently reliably possible to differentiate where potentially unrelated operations, such as reconstructive vascular procedures, are genuinely unrelated, and where they are necessary due to a complication of a previous operation. To reliably capture the full burden of operative intervention for a child, and to reflect what is important to stakeholders, our analysis has therefore captured information about every operation a child has undergone, including operations other than those related directly to the definitive treatment of the child’s Hirschsprung disease. However, as multiple separate OPCS codes are frequently utilised to describe different parts of a single procedure, where multiple surgical procedures are recorded as occurring on the same day, these have been considered as one event.

To understand the extent to which children with HSCR needed learning support, we examined Special Educational Needs recorded in school records by the end of Year 1 (age 6).[24] In England, children are entitled to receive SEN provision if they have “a significantly greater difficulty in learning than the majority of others of the same age, or have a disability which prevents them from making use of facilities generally provided by mainstream schools”. In this study, we include both SEN support (classroom-based support arranged and funded by schools) and Education Health and Care Plans (arranged and funded by local authorities, for children whose needs cannot be met by SEN support) under our SEN grouping.

### Statistical analysis

We described the characteristics of children with and without HSCR, including year of birth, sex, gestational age (<32 weeks, 32-36 weeks, 37+ weeks), birthweight (<1500g, 1500-2500g, 2500g+), small or large for gestation (<10^th^ or >90^th^ percentile of birth weight for gestation derived from national birth weight percentiles),[25] ethnic group (Black, White, Asian, Mixed or Other), maternal age (<=20, 21-30, 31-40 and 40+ years) and quintile of deprivation (IMD of residential postcode). We quantified the number of children with and without HSCR with major cardiac anomalies (ICD-10 codes Q20-Q26) and chromosomal abnormalities including Down Syndrome (ICD-10 codes Q90-93, Q96-Q99, excluding Q93.6).[23] For those with HSCR, we described age at diagnosis.

Our study population was stratified into 5-year birth cohorts (April 2002-March 2007; April 2007-March 2012; April 2012-March 2016) with differing amounts of follow up (up to age 14, age 9 and age 4 respectively, **Supp. Fig. 1**). Since the majority of children with HSCR are diagnosed neonatally, and to ensure comparable time periods for children with and without HSCR, we evaluated the proportion of children who had an admission, a surgical procedure, or death following discharge from their birth episode, up to the full extent of follow up (or death). We then explored the total number of admissions and surgical procedures, and the most frequently occurring reasons for admission according to ICD-10 codes and the high-level chapter of the primary diagnosis code, stratified by sex. We also conducted a stratified analysis for children with and without Down Syndrome.

Analysis of SEN outcomes in Year 1 (age 6) was conducted for the subset of children who were linked to their education record in NPD. We quantified SEN separately for the cohort of children without major cardiac anomalies or chromosomal abnormalities (since many of these children will require extra support at school, irrespective of having HSCR).

For admissions and SEN, we used multivariable generalised linear models to estimate the relative risk of having the outcome, adjusting for sex, ethnicity, maternal age, deprivation, gestational age, and comorbidities. To explore whether the effect of HSCR on SEN differed according to sex or deprivation, we included interaction terms within the models. Analysis was conducted in Stata V17.

### Ethics

Permissions to use linked, de-identified data from Hospital Episode Statistics and the National Pupil Database were granted by the Department of Education (DR200604.02B) and NHS Digital (DARS-NIC-381972). Ethical approval for the ECHILD project was granted by the National Research Ethics Service (17/LO/1494), NHS Health Research Authority Research Ethics Committee (20/EE/0180), and UCL Great Ormond Street Institute of Child Health’s Joint Research and Development Office (20PE06).

## Results

### Cohort characteristics

Of the 11,261,227 live births in England, between April 2002 and March 2020, 3227 (0.03%) had a diagnosis of HSCR coded in their hospital record (corresponding to 1 in 3000 births). Of the 3227 children with HSCR, the majority (65.6%) were diagnosed in the first 28 days of life; an additional 10.4% were diagnosed between 29-90 days; 9.5% were diagnosed age 2 or older, and 4.3% were diagnosed at age 5 or older.

Compared to children without the condition, children with HSCR were more likely to be male, have been born to younger mothers, be living in more deprived areas, preterm, low birth weight, small for gestational age, and have major cardiac anomalies or chromosomal abnormalities (**Table 1**). 9.2% of children with HSCR also had Down Syndrome compared to 0.1% of children with no HSCR (2.0% of children in the cohort with Down Syndrome had HSCR).

**Table 1:** Characteristics of the study cohort (births between April 2002-March 2020)

|  | Total |  | Hirschsprung diagnosis |  | No Hirschsprung diagnosis |  |
| --- | --- | --- | --- | --- | --- | --- |
|  | N | %* | N | %* | N | %* |
| **Total** | 11,261,227 |  | 3227 | 0.03 | 11,258,000 | 99.97 |
| **Sex** |  |  |  |  |  |  |
| Male | 5,766,904 | 51.2 | 2353 | 72.9 | 5,764,551 | 51.2 |
| Female | 5,480,929 | 48.7 | 873 | 27.1 | 5,480,056 | 48.7 |
| Missing |  |  |  |  |  |  |
| **Ethnicity\*** |  |  |  |  |  |  |
| White | 7,657,457 | 76.0 | 2438 | 76.7 | 7,655,019 | 76.0 |
| Black | 533,469 | 5.3 | 185 | 5.8 | 533,284 | 5.3 |
| Asian | 1,076,733 | 10.7 | 274 | 8.6 | 1,076,459 | 10.7 |
| Mixed | 466,551 | 4.6 | 155 | 4.9 | 466,396 | 4.6 |
| Other | 344,087 | 3.4 | 126 | 4.0 | 343,961 | 3.4 |
| Unknown | 1,182,930 | 10.5 | 49 | 1.5 | 1,182,881 | 10.5 |
| **Maternal age\*** |  |  |  |  |  |  |
| <20 | 539,059 | 5.2 | 166 | 5.9 | 538,893 | 5.2 |
| 20-29 | 4,681,567 | 44.9 | 1351 | 47.8 | 4,680,216 | 44.9 |
| 30-39 | 4,821,427 | 46.2 | 1188 | 42.0 | 4,820,239 | 46.2 |
| ≥40 | 394,848 | 3.8 | 122 | 4.3 | 394,726 | 3.8 |
| Missing | 824,326 | 7.3 | 400 | 12.4 | 823,926 | 7.3 |
| **Quintile of deprivation (IMD)\*** |  |  |  |  |  |  |
| Most deprived | 2,231,324 | 25.6 | 936 | 29.4 | 2,230,388 | 25.6 |
| 2 | 1,776,098 | 20.4 | 645 | 20.2 | 1,775,453 | 20.4 |
| 3 | 1,489,401 | 17.1 | 554 | 17.4 | 1,488,847 | 17.1 |
| 4 | 1,316,385 | 15.1 | 477 | 15.0 | 1,315,908 | 15.1 |
| Least deprived | 1,888,650 | 21.7 | 576 | 18.1 | 1,888,074 | 21.7 |
| Missing | 2,512,370 | 22.3 | 38 | 1.2 | 2,515,332 | 22.3 |
| **Gestational age (weeks)\*** |  |  |  |  |  |  |
| <32 | 104,176 | 1.2 | 79 | 3.3 | 104,097 | 1.2 |
| 32-36 | 534,113 | 6.0 | 248 | 10.4 | 533,865 | 6.0 |
| 37+ | 8,248,788 | 92.8 | 2064 | 86.3 | 8,246,724 | 92.8 |
| Missing | 2,374,150 | 21.1 | 836 | 25.9 | 2,373,314 | 21.1 |
| **Birth weight (g)\*** |  |  |  |  |  |  |
| <1500 | 93895 | 1.0 | 63 | 2.5 | 93,832 | 1.0 |
| 1500-2500 | 554010 | 5.9 | 238 | 9.5 | 553,772 | 5.9 |
| >2500 | 8727676 | 93.1 | 2214 | 88.0 | 8,725,462 | 93.1 |
| Missing | 1885646 | 16.7 | 712 | 22.1 | 1,884,934 | 16.7 |
| **Size for gestation\*** |  |  |  |  |  |  |
| Small (<10<sup>th</sup> percentile) | 605,184 | 7.0 | 177 | 7.6 | 605,007 | 7.0 |
| Normal | 7,304,306 | 84.5 | 1965 | 84.4 | 7,302,341 | 84.5 |
| Large (>90<sup>th</sup> percentile) | 739,402 | 8.5 | 187 | 8.0 | 739,215 | 8.5 |
| Missing | 2,612,335 | 23.2 | 898 | 27.8 | 2,611,437 | 23.2 |
| **Comorbidities** |  |  |  |  |  |  |
| Major cardiac anomalies (any) | 156,507 | 1.4 | 493 | 15.3 | 156,014 | 1.4 |
| ...of cardiac chambers and connections | 10608 | 0.1 | 10590 | 0.1 | 18 | 0.6 |
| ...of cardiac septa | 95624 | 0.8 | 95218 | 0.8 | 406 | 12.6 |
| ...of pulmonary and tricuspid valves | 16245 | 0.1 | 16190 | 0.1 | 55 | 1.7 |
| ...Other congenital malformations of heart | 14392 | 0.1 | 14352 | 0.1 | 40 | 1.2 |
| ...of great arteries | 22155 | 0.2 | 22089 | 0.2 | 66 | 2 |
| ...of great veins | 93271 | 0.8 | 92993 | 0.8 | 278 | 8.6 |
| Chromosomal abnormalities (any) | 31,357 | 0.3 | 373 | 11.6 | 30,984 | 0.3 |
| ...Down syndrome | 13,230 | 0.1 | 271 | 9.2 | 12,959 | 0.1 |
| ...Edwards syndrome and Patau syndrome | 1571 | 0.0 | ** | 0.0 | <10 | <0.3 |
| ...Other trisomies and partial trisomies of the autosomes, NEC | 3216 | 0.0 | 3194 | 0.0 | 22 | 0.7 |
| ...Monosomies and deletions from the autosomes, not elsewhere classified | 8938 | 0.1 | 8889 | 0.1 | 49 | 1.5 |
| ...Turner syndrome | 1256 | 0.0 | ** | 0.0 | <10 | <0.3 |
| ...Other sex chromosome abnormalities, female phenotype, NEC | 649 | 0.0 | ** | 0.0 | <10 | <0.3 |
| ...Other sex chromosome abnormalities, male phenotype, NEC | 1280 | 0.0 | ** | 0.0 | <10 | <0.3 |
| Primary or secondary pulmonary hypertension | 7717 | 0.1 | 7682 | 0.1 | 35 | 1.1 |

\* Percentages are of complete records. NEC = Not Elsewhere Classified.
\*\* Cell sizes <10 and those which would allow small cell sizes to be derived have been suppressed due to statistical disclosure control.

### Rates of hospital admissions and surgical procedures

Children with HSCR were more likely than those without to be admitted for any reason than those without HSCR: 95.0% of children with HSCR were admitted at least once by age 4 compared with 40.2% of those without (**Table 2**). Children with HSCR had a median of 6 admissions (interquartile range 3-12) by age 4, compared with a median of 0 admissions (interquartile range 0-1) for those without HSCR. Over a quarter of children with HSCR had 10 or more admissions by age 4 (**Supp. Table 2**).

**Table 2:** Number and percentage of children with and without Hirschsprung disease admitted to hospital, by age and sex.

|  | No Hirschsprung diagnosis |  |  | Hirschsprung diagnosis |  |  |
| --- | --- | --- | --- | --- | --- | --- |
|  | Total | Children admitted |  | Total | Children admitted |  |
|  |  | N | % |  | N | % |
| All |  |  |  |  |  |  |
| Admission aged 0-4 years |  |  |  |  |  |  |
| Cohort 1: born between 2002/03 and 2016/17 | 9,435,790 | 3,790,361 | 40.2 | 2700 | 2,566 | 95.0 |
| Admission aged 5-9 years |  |  |  |  |  |  |
| Cohort 2: born between 2002/03 and 2011/12 | 6,200,844 | 1,210,703 | 19.5 | 1768 | 920 | 52.0 |
| Admission aged 10-14 years |  |  |  |  |  |  |
| Cohort 3: born between 2002/03 and 2006/07 | 2,910,568 | 427,511 | 14.7 | 855 | 320 | 37.4 |
| Girls |  |  |  |  |  |  |
| Admission aged 0-4 years |  |  |  |  |  |  |
| Cohort 1: born between 2002/03 and 2016/17 | 4593676 | 1,672,581 | 36.4 | 733 | 680 | 92.8 |
| Admission aged 5-9 years |  |  |  |  |  |  |
| Cohort 2: born between 2002/03 and 2011/12 | 3018246 | 533,228 | 17.7 | 469 | 234 | 49.9 |
| Admission aged 10-14 years |  |  |  |  |  |  |
| Cohort 3: born between 2002/03 and 2006/07 | 1418725 | 197,343 | 13.9 | 225 | 73 | 32.4 |
| Boys |  |  |  |  |  |  |
| Admission aged 0-4 years |  |  |  |  |  |  |
| Cohort 1: born between 2002/03 and 2016/17 | 4829738 | 2,117,780 | 43.8 | 1966 | 1,886 | 95.9 |
| Admission aged 5-9 years |  |  |  |  |  |  |
| Cohort 2: born between 2002/03 and 2011/12 | 3170930 | 677,475 | 21.4 | 1299 | 686 | 52.8 |
| Admission aged 10-14 years |  |  |  |  |  |  |
| Cohort 3: born between 2002/03 and 2006/07 | 1487034 | 230,168 | 15.5 | 630 | 247 | 39.2 |

Admission rates decreased with age but were still substantially higher by age 14 in children with HSCR compared to those without. Boys were more likely to be admitted than girls. For children with Down Syndrome, admission rates were still higher amongst the group with HSCR: 95.5% of those with HSCR were admitted by age 4 compared with 85.8% of those without HSCR; 65.0% versus 60.7% were admitted between ages 5-9, and 64.1% versus 48.7% were admitted between ages 10-14 (**Supp. Table 3**).

Of children with HSCR, 81.4% had 2 or more surgical procedures between ages 0-4, compared to 2.9% in children without HSCR (**Table 3**, **Supp. Table 4**). Overall numbers of surgical procedures remained high at ages 5-9 and 10-14, with differences between children with and without HSCR persisting. The most commonly recorded surgical procedures were H41.2 (Peranal excision of lesion of rectum; 73.7%) and H41.8 (Other operations on rectum through anus; 61.1%) for ages 0-4, H56.8 (Other operations on anus; 6.3%) for ages 5-9, and G45.1 (Fibreoptic endoscopic examination of upper gastrointestinal tract and biopsy of lesion of upper gastrointestinal tract; 3.6%) for ages 10-14 (**Supp. Table 5**).

**Table 3:** Number of surgical procedures for children with and without Hirschsprung disease admitted to hospital, by age and sex.

| N procedures | All |  |  |  | Boys |  |  |  | Girls |  |  |  |
| --- | --- | --- | --- | --- | --- | --- | --- | --- | --- | --- | --- | --- |
|  | No HSCR |  | HSCR |  | No HSCR |  | HSCR |  | No HSCR |  | HSCR |  |
|  | N | % | N | % | N | % | N | % | N | % | N | % |
| **Age 0-4** | 9,435,790 |  | 2700 |  | 4,829,738 |  | 1966 |  | 4,593,676 |  | 733 |  |
| 1 | 919,105 | 9.7 | 300 | 11.1 | 551,895 | 11.4 | 191 | 9.7 | 367,073 | 8.0 | 109 | 14.9 |
| 2-3 | 206,444 | 2.2 | 976 | 36.1 | 130,867 | 2.7 | 709 | 36.1 | 75,555 | 1.6 | 266 | 36.3 |
| 4-5 | 33,039 | 0.4 | 582 | 21.6 | 19,624 | 0.4 | 437 | 22.2 | 13,411 | 0.3 | 145 | 19.8 |
| 6-9 | 17,343 | 0.2 | 449 | 16.6 | 10,062 | 0.2 | 356 | 18.1 | 7,279 | 0.2 | 93 | 12.7 |
| 10+ | 13,265 | 0.1 | 191 | 7.1 | 7,421 | 0.2 | 144 | 7.3 | 5,843 | 0.1 | 47 | 6.4 |
| **Age 5-9** | 6,200,844 |  | 1768 |  | 3,170,930 |  | 1299 |  | 3,018,246 |  | 469 |  |
| 1 | 1,023,620 | 16.5 | 175 | 9.9 | 583,739 | 18.4 | 111 | 8.5 | 439,782 | 14.6 | 64 | 13.6 |
| 2-3 | 337,656 | 5.4 | 563 | 31.8 | 207,774 | 6.6 | 416 | 32.0 | 129,863 | 4.3 | 147 | 31.3 |
| 4-5 | 47,712 | 0.8 | 377 | 21.3 | 28,970 | 0.9 | 271 | 20.9 | 18,738 | 0.6 | 106 | 22.6 |
| 6-9 | 21,848 | 0.4 | 338 | 19.1 | 12,757 | 0.4 | 269 | 20.7 | 9,089 | 0.3 | 69 | 14.7 |
| 10+ | 17,723 | 0.3 | 227 | 12.8 | 10,004 | 0.3 | 178 | 13.7 | 7,718 | 0.3 | 49 | 10.4 |
| **Age 10-14** | 2,910,568 |  | 855 |  | 1,487,034 |  | 630 |  | 1,418,725 |  | 225 |  |
| 1 | 536,308 | 8.6 | 88 | 10.3 | 299,974 | 20.2 | 55 | 8.7 | 236,270 | 16.7 | 33 | 14.7 |
| 2-3 | 225,927 | 3.6 | 249 | 29.1 | 136,539 | 9.2 | 178 | 28.3 | 89,370 | 6.3 | 71 | 31.6 |
| 4-5 | 36,801 | 0.6 | 171 | 20.0 | 22,388 | 1.5 | 137 | 21.7 | 14,409 | 1.0 | 34 | 15.1 |
| 6-9 | 16,534 | 0.3 | 173 | 20.2 | 9,705 | 0.7 | 137 | 21.7 | 6,827 | 0.5 | 36 | 16.0 |
| 10+ | 13,124 | 0.2 | 127 | 14.9 | 7,412 | 0.5 | 94 | 14.9 | 5,711 | 0.4 | 33 | 14.7 |

In analyses controlling for key risk factors (sex, ethnicity, deprivation, gestational age at birth, comorbidities), children with HSCR were 42% more likely to have been admitted between ages 0-4 (Incidence Rate Ratio [IRR] 1.47; 95% CI 1.40-1.53), 40% more likely to have been admitted between ages 5-9 (IRR 1.40; 95% CI 1.29-1.51), and 35% more likely to have been admitted between ages 10-14 (IRR 1.35; 95% CI 1.17-1.55), compared to those without HSCR.

### Reasons for admission

Across all age groups, children with HSCR were more likely than those without to be admitted with diagnoses falling under virtually all ICD-10 diagnosis chapters (**Supp. Fig. 2**, **Supp. Table 6**). The largest differences in admission rates were seen in diagnoses relating to diseases of the digestive system. Reasons for admission were relatively similar for girls and boys (**Supp. Tables 7-8** for ages 0-4 and ages 5-9; numbers were too small to stratify for the age 10-14 cohort).

The most frequently occurring diagnoses in admissions within the HSCR group across all age groups and sexes was constipation (affecting ∼35% of children; **Fig. 1**, **Supp. Table 9**), followed by gastroenteritis and intestinal infections. Around 15% of children across all age groups were readmitted for nausea and vomiting, and 8-10% were readmitted for abdominal pain. Across all age groups, 8-11% of children were readmitted with a primary diagnosis of “flatulence and related conditions” (ICD-10 code R14, which covers various digestive and abdominal symptoms, including an inability to pass flatus [i.e. functional sphincter achalasia] as well as bloating, belching, and gas pain).

**Figure 1:**
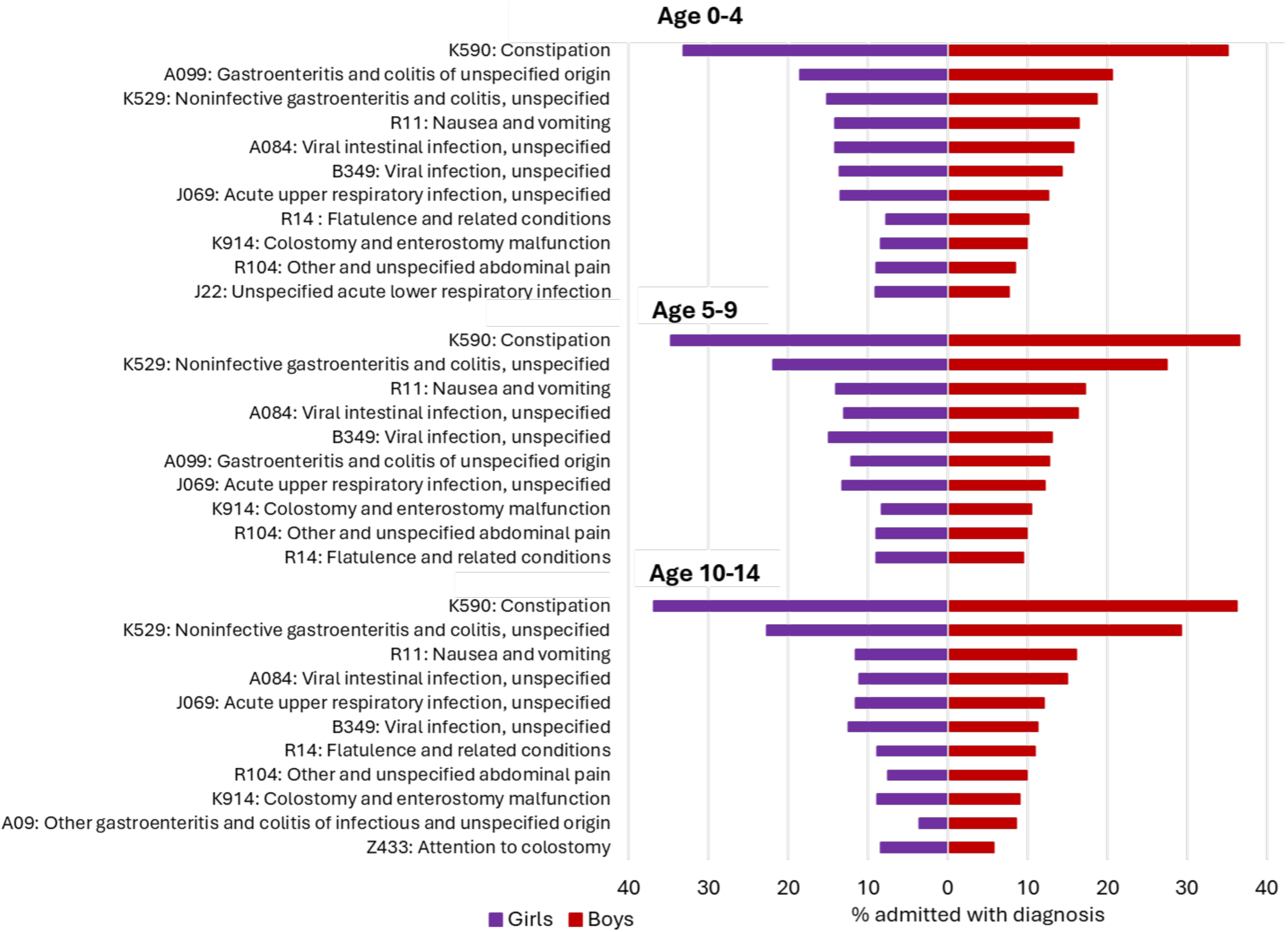
Most frequently occurring ICD 10 codes in primary diagnosis fields for children with Hirschsprung, by age and sex.

### Mortality

Mortality rates were substantially higher in children with HSCR: 86/2700 (3.2%) of children with HSCR had died by age 4 compared with 0.5% in children without HSCR; 10/1768 (0.6%) of children with HSCR died between ages 5-9 compared to 0.03% of children without HSCR. Excluding those with chromosomal abnormalities or major cardiac anomalies, 39/2217 (1.8%) children with HSCR had died by age 4 compared with 35804/9294385 (0.4%) of children without HSCR. Numbers for deaths between ages 10-14, or deaths aged 5-9 for those without comorbidities were too low to report.

### Special Educational Needs at school age

Within the 8,046,292 (71.5%) of children who were linked to a school record in Year 1 (age 6) NPD, 892/2025 (44.0%) of those with HSCR had recorded Special Educational Needs compared with 1,213,133/6,777,687 (17.9%) of those without HSCR. For the cohort of children without chromosomal anomalies, 680/1807 (37.6%) of those with HSCR had recorded Special Educational Needs compared with 1195761/6757836 (17.7%) of those without HSCR. In adjusted analysis controlling for sex, ethnicity, maternal age, deprivation, gestational age, and comorbidities, children with HSCR were 34% more likely to have recorded SEN by Year 1 compared to those without HSCR (IRR 1.34; 95% CI 1.24-1.45). Interaction terms indicated that there was no difference in the effect of HSCR on SEN by quintile of deprivation. However, the impact of HSCR on SEN was greater for females (IRR 1.57; 95% CI 1.33-1.85) compared to males (IRR 1.36; 95% CI 1.25-1.49; p-value for interaction <0.0001).

## Discussion

Our study aimed to fill a gap in evidence on longitudinal outcomes for children with HSCR at a population level, describing characteristics and outcomes of children with HSCR including need for hospitalisation (up to 14 years of age) and educational needs. To the best of our knowledge, this represents the first time that a national database spanning multiple decades has been used to evaluate the frequency of hospital admissions in a HSCR cohort, as well as the specific health problems precipitating admissions. Our study highlights that irrespective of common comorbidities such as Down Syndrome, children with HSCR have a significantly higher mortality rate and are admitted to hospital frequently, and more often than children without HSCR, at least up to age 14. Importantly, these admissions are often for constipation, enterocolitis and abdominal pain, suggesting ongoing severe sequelae despite surgical intervention. Our data also highlight shifts in admission of children with HSCR for specific diagnosis according to age. Interestingly, we observed that ∼10% of children with HSCR were admitted with diagnosis codes for flatulence and related conditions across all age groups. Although this ICD-10 code covers various digestive and abdominal symptoms, including bloating, belching, and gas pain, this finding points towards ongoing issues surrounding bowel habits which potentially impact early schooling (through disruption caused by issues with toileting) and quality-of-life during adolescence.[26, 27] These findings support previously published work that showed 49% of children with HSCR scored one standard deviation below the reference population in terms of quality-of-life measures, and 42% of children with HSCR reporting significantly reduced psychological well-being.[28]

While surgery for HSCR is lifesaving, patients often display significant downstream issues affecting their quality of life.[29–32] The aetiology of these ongoing symptoms is currently incompletely understood despite substantial efforts.[23, 33–38] For example, the NETS^2HD^ UK-based study, a prospective cohort study conducted from 2010 to 2012, detailed specific outcomes that could be tracked following surgical intervention to better inform surgical practice.[28] This included data from 28 UK and Irish paediatric surgical centres. The related NETS^1HD^ study developed a core outcome set to standardise surgical success metrics, such as ‘long-term faecal incontinence’ and a ‘need for a permanent stoma’.[12] However, this has not yet been fully implemented or evaluated, and there is currently no clear consensus on the most effective surgical technique, further highlighting the importance of evaluating long-term functional results.

Our findings support evidence from several other studies that have conclusively demonstrated ongoing HSCR-related symptoms persisting post-surgery, into adult life.[39, 40] NETS^2HD^, which examined outcomes in children with HSCR at five to eight years of age in the UK, found that 44% of children in this cohort had undergone at least one unplanned reoperation, with 27% classified as major or complex procedures.[28] However, previous studies have often been focussed on specific outcomes, such as diarrhoea and constipation, with many studies relying on patient self-reporting QoL outcomes.[29, 41, 42] [43, 44]

In addition to increased mortality and ongoing health issues, we also found that children with HSCR were significantly more likely to have recorded Special Educational Needs at the start of compulsory schooling. A recent systematic review has outlined reductions in school functioning in children with HSCR.[45] This review particularly highlighted issues surrounding peer relationships, though other indicators such as school performance, teacher relationships, and need for special education services varied widely and were less consistently measured, pointing to the need for large longitudinal studies with empirical metrics to better understand and address the educational needs in HSCR. While it is well established that HSCR can be associated with chromosomal abnormalities (e.g., Trisomy 21) our study shows that even after controlling for these conditions, children with HSCR remained 34% more likely to have recorded SEN by Year 1 compared to those without HSCR.[46–48] More research is needed to understand the extent to which this is explained by organic cognitive issues, ongoing needs to manage their condition, or increased hospitalisations which impact on learning. Interestingly, a Swedish study found that the adult HSCR population (aged 16-49) achieved comparable earning potential and educational qualifications compared to age and sex-matched individuals in the general population.[49] This may suggest that HSCR does not itself impact on learning potential, but perhaps that recorded SEN in the current study represents the impact of extended educational absences due to frequent hospital admissions or the need for practical assistance within school to deal with on ongoing symptoms. Of note, the impact of HSCR on educational outcomes is highly likely to be country specific, as the ability of national education sectors to incorporate these HSCR into school, and keep them in education is likely to vary significantly country to country. However, regardless of cause, our data highlight a critical ongoing need for children with HSCR and their families, particularly during formative years.

A major strength of our study is the population-level coverage of our data, capturing ∼97% of births in NHS hospitals in England. Our findings are comparable with other literature using administrative health records to analyse outcomes of HSCR: a previous study using HES found that 85.3% of those with a diagnosis had an operation within the first year of life (in our study, this was 87.4%).[23] The longitudinal nature of the dataset allowed us to evaluate longer-term outcomes and to link with outcomes outside of health, in order to start to understand the experiences of children with HSCR in school. We specifically focussed on children aged up to 14, before the transition from paediatric to adult services at the age of 16. Difficulties in management of disease are often experienced at this transition period, and future work will look separately at older adolescents, including the impact that HSCR has on their mental health.[50, 51]

A limitation of our study is the nature of recording of ICD-10 diagnosis codes and OPCS procedure codes; these codes are used primarily for billing purposes and there may be some misclassification. We relied on diagnosis dates as recorded in inpatient records. However, diagnoses may not always be made as an inpatient (e.g., if a biopsy is returned after discharge) and in this case, would not be coded until a later hospital admission (in which case the true diagnosis date would have been earlier than we identified). A further limitation is that we were not able to distinguish between different types or severities of HSCR, and the genetic basis for HSCR in individual patients is not recorded. Nevertheless, our study provides an improved understanding of HSCR burden highlighting the real-world impact on children and their families/support groups.

Clinical management of HSCR is primarily based on the success of early surgical interventions. However, despite well-established treatment regimens for HSCR, children with this condition remain at higher risk of chronic clinical complications and potential developmental delays. Greater detail in these areas may guide targeted interventions, tailored multidisciplinary follow-up and potentially earlier intervention for these children. Additionally, our data highlights the need for ongoing support of children with HSCR throughout childhood and adolescence, providing healthcare systems and carers with crucial information which can be used to anticipate and plan long-term needs and resources. Indeed, evidence suggests that families with a new diagnosis of HSCR often feel under-informed as to the potential impacts of the condition on their child’s development.[43] For example, newly diagnosed patients with relatives affected by HSCR report better outcomes, possibly suggesting that better information is linked to higher quality of life.[44]

Moreover, our study highlights that while surgical intervention is essential, it does not provide complete resolution of the condition, with outcomes varying widely. Hence, there is a critical need for novel approaches to improve treatment success and quality-of-life for people living with HSCR. In addition, further research is also needed to understand the impact of HSCR and its treatment at the transition from paediatric to adult services, and to evaluate child development, educational attainment, and other psychosocial factors including mental health.

## Data Availability

All data produced in the present work are contained in the manuscript

## Acknowledgements

We gratefully acknowledge all children and families whose de-identified data were used in this research. The ECHILD Database uses data from the Department for Education (DfE). The DfE does not accept responsibility for any inferences or conclusions derived by the authors. This work contains statistical data from ONS which is Crown Copyright. The use of the ONS statistical data in this work does not imply the endorsement of the ONS in relation to the interpretation or analysis of the statistical data. This work uses research datasets which may not exactly reproduce National Statistics aggregates. Evidence from this research contributes to the NIHR Children and Families Policy Research Unit but was not commissioned by the NIHR Policy Research Programme.

Part of this research was conducted at UCL Great Ormond Street Institute of Child Health and supported by the NIHR Great Ormond Street Hospital Biomedical Research Centre. Views expressed in this manuscript are solely those of the authors and not necessarily those of the NHS, the NIHR or the Department of Health.

## Contributions

KH: conceptualisation, funding acquisition, data curation, formal analysis, investigation, writing original draft, writing – review editing. BJ: conceptualisation, writing original draft, writing – review editing. BA: data analysis review, writing – review editing. JD: data analysis review, writing – review editing. AT: funding acquisition, writing – review editing. PWA: conceptualisation, funding acquisition, writing – review editing. PDC: funding acquisition, writing – review editing. CMC: conceptualisation, funding acquisition, writing original draft, writing – review editing

## Supplementary material

**Supplementary Figure 1:**
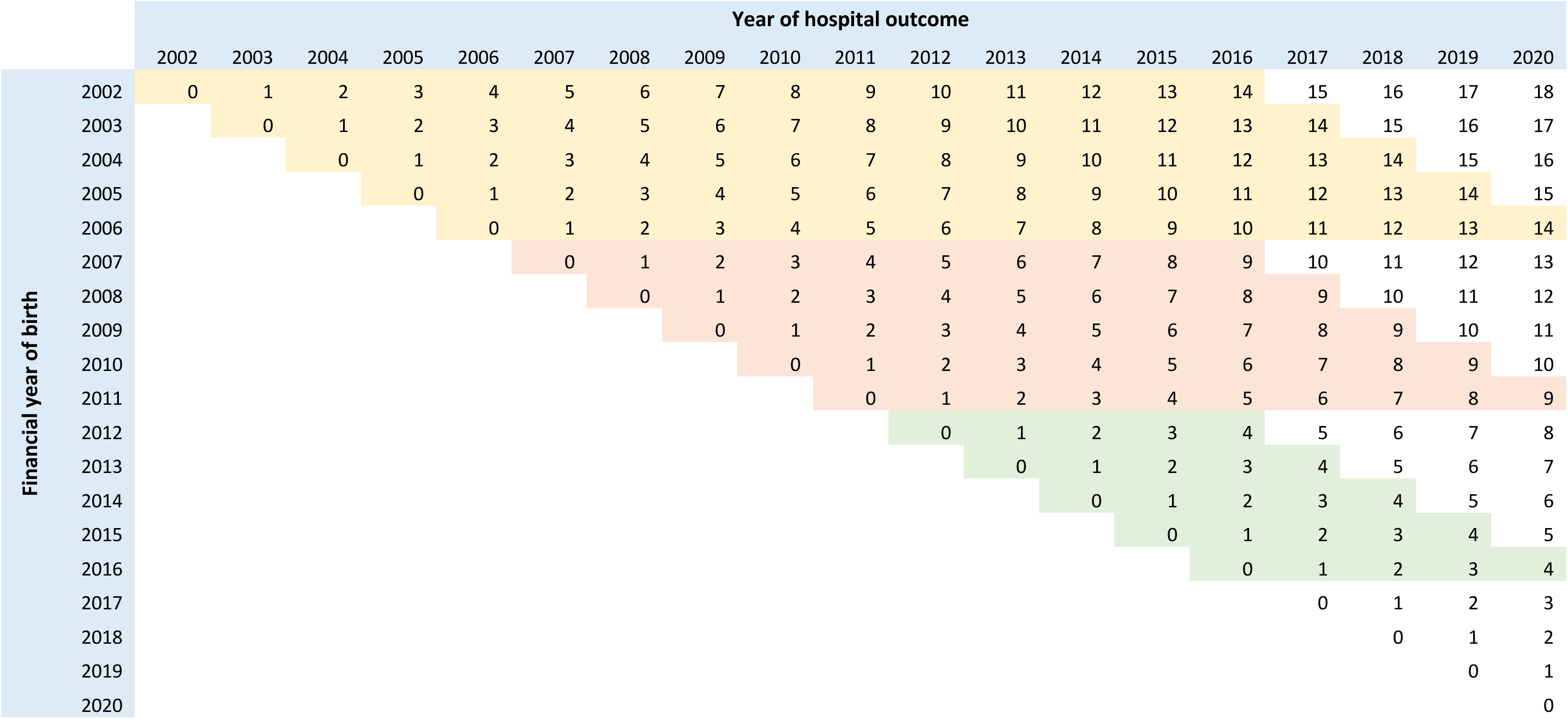
Age at hospital outcome (in boxes) for the different cohorts. Orange shaded sections indicate the earliest cohort, born between April 2002 and March 2007 (followed up to age 14). Red shaded sections indicate the middle cohort, born between April 2007 and March 2012 (followed up to age 9). Green shaded boxes indicate the latest cohort, born between April 2012 and March 2017 (follow up to age 4).

**Supplementary Figure 2:**
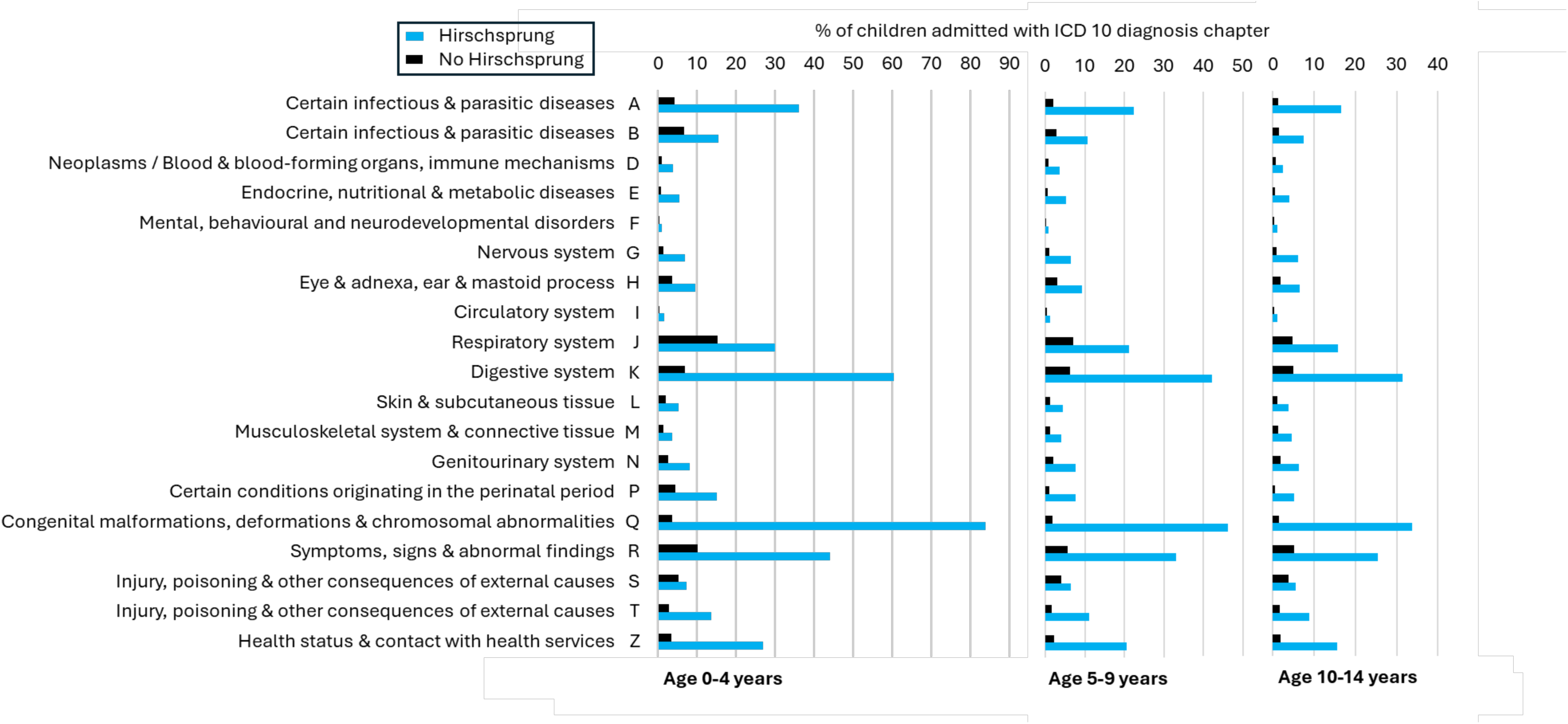
Percentage of children with admissions according to ICD-10 chapter headings for children with and without Hirschsprung disease, by age group

**Supplementary Table 1:** OPCS codes excluded from analysis of procedures/operations.

| OPCS Code | Description |
| --- | --- |
| Any U code | Diagnostic imaging, testing and rehabilitation |
| Any Y code | Subsidiary classification of methods of operation |
| Any Z code | Subsidiary classification of sites of operation |
| A55.9 | Diagnostic spinal puncture, Unspecified |
| A84.1 | Electroencephalography NEC |
| A84.4 | Evoked potential recording |
| C87.8 | Evaluation of retina, other specified |
| E08.8 | Other operations on internal nose, other specified |
| E85.1 | Invasive ventilation |
| E85.2 | Non-invasive ventilation NEC |
| E85.4 | Bag valve mask ventilation |
| E85.6 | Continuous positive airway pressure |
| G21.4 | Intubation of oesophagus NEC |
| G47.5 | Insertion of nasogastric tube |
| G47.8 | Intubation of stomach, other specified |
| G47.9 | Intubation of stomach, unspecified |
| H62.5 | Irrigation of bowel NEC |
| L26.4 | Aortography |
| L91.1 | Open insertion of central venous catheter |
| L91.2 | Insertion of central venous catheter NEC |
| L91.3 | Attention to central venous catheter NEC |
| L91.4 | Removal of central venous catheter |
| L91.5 | Insertion of tunnelled venous catheter |
| L92.3 | Thrombolysis of access catheter |
| L99.7 | Percutaneous transluminal peripheral insertion of central catheter |
| M47.3 | Removal of urethral catheter from bladder |
| M47.9 | Urethral catheterisation of bladder, unspecified |
| O15.2 | Insertion of umbilical venous catheter |
| O15.3 | Insertion of umbilical arterial catheter |
| O30.1 | Hepatic flexure |
| O48.1 | Prone positioning of patient |
| S12.8 | Phototherapy to skin, other specified |
| S12.9 | Phototherapy to skin, unspecified |
| S57.5 | Attention to dressing of skin NEC |
| T20.3 | Primary repair of inguinal hernia using sutures |
| X29.2 | Continuous intravenous infusion of therapeutic substance NEC |
| X32.3 | Exchange of plasma (2-9) |
| X36.8 | Blood withdrawal, other specified |
| X36.9 | Blood withdrawal, unspecified |
| X39.8 | Other route of administration of therapeutic substance, other specified |
| X40.6 | Continuous ambulatory peritoneal dialysis |
| X50.1 | Direct current cardioversion |
| X50.3 | Advanced cardiac pulmonary resuscitation |
| X52.2 | High flow nasal oxygen therapy |
| X52.8 | Oxygen therapy, other specified |
| X56.1 | Nasotracheal intubation |
| X56.2 | Endotracheal intubation |
| X56.8 | Intubation of trachea, other specified |
| X82.1 | Pulmonary arterial hypertension drugs Band 1 |
| X84.1 | Medical gases Band 1 |
| X85.1 | Torsion dystonias and other involuntary movements drugs Band 1 |
| X86.5 | Respiratory syncytial virus prevention drugs Band 1 |
| X90.4 | Intravenous nutrition Band 1 |
| X93.1 | Subfoveal choroidal neovascularisation drugs Band 1 |
| X96.1 | Immunoglobulins Band 1 |

NEC = Not Elsewhere Classified

**Supplementary Table 2:** Number of admissions by age, sex and whether the child had Down Syndrome, for children with and without HSCR.

| N admissions | All |  |  |  | Boys |  |  |  | Girls |  |  |  | No Down Syndrome |  |  |  | Down Syndrome |  |  |  |
| --- | --- | --- | --- | --- | --- | --- | --- | --- | --- | --- | --- | --- | --- | --- | --- | --- | --- | --- | --- | --- |
|  | No HSCR |  | HSCR |  | No HSCR |  | HSCR |  | No HSCR |  | HSCR |  | No HSCR |  | HSCR |  | No HSCR |  | HSCR |  |
|  | N | % | N | % | N | % | N | % | N | % | N | % | N | % | N | % | N | % | N | % |
| Age 0-4 | 9,435,790 |  | 2,700 |  | 4,829,738 |  | 1,966 |  | 4,593,676 |  | 733 |  | 9,424,933 |  | 2,477 |  | 10,857 |  | 223 |  |
| 0 | 5,645,429 | 59.8 | 134 | 5.0 | 2,711,958 | 56.2 | 80 | 4.1 | 2,921,095 | 63.6 | 53 | 7.2 | 5,643,883 | 59.9 | 124 | 5.0 | 1,546 | 14.2 | 10 | 4.5 |
| 1 | 2,136,681 | 22.6 | 245 | 9.1 | 1,142,567 | 23.7 | 156 | 7.9 | 994,114 | 21.6 | 89 | 12.1 | 2,135,268 | 22.7 | 237 | 9.6 | 1,413 | 13.0 | <10* |  |
| 2-3 | 1,207,622 | 12.8 | 558 | 20.7 | 701,683 | 14.5 | 405 | 20.6 | 505,939 | 11.0 | 153 | 20.9 | 1,205,119 | 12.8 | 529 | 21.4 | 2,503 | 23.1 | 29 | 13.0 |
| 4-5 | 263,989 | 2.8 | 478 | 17.7 | 161,930 | 3.4 | 346 | 17.6 | 102,059 | 2.2 | 132 | 18.0 | 262,186 | 2.8 | 447 | 18.0 | 1,803 | 16.6 | 31 | 13.9 |
| 6-9 | 120,614 | 1.3 | 571 | 21.1 | 74,992 | 1.6 | 445 | 22.6 | 45,622 | 1.0 | 126 | 17.2 | 118,826 | 1.3 | 509 | 20.5 | 1,788 | 16.5 | 62 | 27.8 |
| 10+ | 61,455 | 0.7 | 714 | 26.4 | 36,608 | 0.8 | 534 | 27.2 | 24,847 | 0.5 | 180 | 24.6 | 59,651 | 0.6 | 631 | 25.5 | 1,804 | 16.6 | 83 | 37.2 |
| Age 5-9 | 6,200,844 |  | 1,768 |  | 3,170,930 |  | 1,299 |  | 3,018,246 |  | 469 |  | 6,193,771 |  | 1,631 |  | 7,073 |  | 137 |  |
| 0 | 4,990,141 | 80.5 | 848 | 48.0 | 2,493,455 | 78.6 | 613 | 47.2 | 2,485,018 | 82.3 | 235 | 50.1 | 4,987,362 | 80.5 | 800 | 49.0 | 2,779 | 39.3 | 48 | 35.0 |
| 1 | 841,908 | 13.6 | 312 | 17.6 | 466,502 | 14.7 | 239 | 18.4 | 375,406 | 12.4 | 73 | 15.6 | 840,422 | 13.6 | 298 | 18.3 | 1,486 | 21.0 | 14 | 10.2 |
| 2-3 | 291,932 | 4.7 | 287 | 16.2 | 167,635 | 5.3 | 209 | 16.1 | 124,297 | 4.1 | 78 | 16.6 | 290,351 | 4.7 | 247 | 15.1 | 1,581 | 22.4 | 40 | 29.2 |
| 4-5 | 42,220 | 0.7 | 95 | 5.4 | 24,249 | 0.8 | 76 | 5.9 | 17,971 | 0.6 | 19 | 4.1 | 41,625 | 0.7 | 85 | 5.2 | 595 | 8.4 | 10 | 7.3 |
| 6-9 | 19,171 | 0.3 | 117 | 6.6 | 10,686 | 0.3 | 85 | 6.5 | 8,485 | 0.3 | 32 | 6.8 | 18,814 | 0.3 | 106 | 6.5 | 357 | 5.0 | 11 | 8.0 |
| 10+ | 15,472 | 0.2 | 109 | 6.2 | 8,403 | 0.3 | 77 | 5.9 | 7,069 | 0.2 | 32 | 6.8 | 15,197 | 0.2 | 95 | 5.8 | 275 | 3.9 | 14 | 10.2 |
| Age 10-14 | 2,910,568 |  | 855 |  | 1,487,034 |  | 630 |  | 1,418,725 |  | 225 |  | 2,907,234 |  | 791 |  | 3,334 |  | 64 |  |
| 0 | 2,483,057 | 85.3 | 535 | 62.6 | 1,256,866 | 84.5 | 383 | 60.8 | 1,221,382 | 86.1 | 152 | 67.6 | 2,481,345 | 85.4 | 512 | 64.7 | 1,712 | 51.3 | 23 | 35.9 |
| 1 | 295,575 | 10.2 | 139 | 16.3 | 161,083 | 10.8 | 106 | 16.8 | 134,492 | 9.5 | 33 | 14.7 | 294,899 | 10.1 | 123 | 15.5 | 676 | 20.3 | 16 | 25.0 |
| 2-3 | 100,004 | 3.4 | 94 | 11.0 | 53,227 | 3.6 | 78 | 12.4 | 46,777 | 3.3 | 16 | 7.1 | 99,467 | 3.4 | 81 | 10.2 | 537 | 16.1 | 13 | 20.3 |
| 4-5 | 16,506 | 0.6 | 38 | 4.4 | 8,142 | 0.5 | 33 | 5.2 | 8,364 | 0.6 | 24** | 10.7 | 16,311 | 0.6 | 31 | 3.9 | 195 | 5.8 | 12** | 18.8 |
| 6-9 | 8,304 | 0.3 | 25 | 2.9 | 4,059 | 0.3 | 16 | 2.5 | 4,245 | 0.3 |  | 0.0 | 8,192 | 0.3 | 22 | 2.8 | 112 | 3.4 |  |  |
| 10+ | 7,122 | 0.2 | 24 | 2.8 | 3,657 | 0.2 | 14 | 2.2 | 3,465 | 0.2 |  | 0.0 | 7,020 | 0.2 | 22 | 2.8 | 102 | 3.1 |  |  |

\* Number suppressed due to small cell size; \*\* 4+ admissions have been grouped due to small numbers

**Supplementary Table 3:** Number and percentage of children with and without Hirschsprung disease admitted to hospital, by age and whether the child had Down Syndrome.

|  | No Hirschsprung diagnosis |  |  | Hirschsprung diagnosis |  |  |
| --- | --- | --- | --- | --- | --- | --- |
|  | Total | Children admitted |  | Total | Children admitted |  |
|  |  | N | % |  | N | % |
| Children without Down Syndrome |  |  |  |  |  |  |
| Admission aged 0-4 years |  |  |  |  |  |  |
| Cohort 1: born between 2002/03 and 2016/17 | 9,424,933 | 3,781,050 | 40.1 | 2,477 | 2,353 | 95.0 |
| Admission aged 5-9 years |  |  |  |  |  |  |
| Cohort 2: born between 2002/03 and 2011/12 | 6,193,771 | 1,206,409 | 19.5 | 1,631 | 831 | 51.0 |
| Admission aged 10-14 years |  |  |  |  |  |  |
| Cohort 3: born between 2002/03 and 2006/07 | 2,907,234 | 425,889 | 14.6 | 791 | 279 | 35.3 |
| Children with Down Syndrome |  |  |  |  |  |  |
| Admission aged 0-4 years |  |  |  |  |  |  |
| Cohort 1: born between 2002/03 and 2016/17 | 10,857 | 9,311 | 85.8 | 223 | 213 | 95.5 |
| Admission aged 5-9 years |  |  |  |  |  |  |
| Cohort 2: born between 2002/03 and 2011/12 | 7,073 | 4,294 | 60.7 | 137 | 89 | 65.0 |
| Admission aged 10-14 years |  |  |  |  |  |  |
| Cohort 3: born between 2002/03 and 2006/07 | 3,334 | 1,622 | 48.7 | 64 | 41 | 64.1 |

**Supplementary Table 4:** Number of surgical procedures by whether the child had Down Syndrome, for children with and without HSCR.

| N<br>procedures | No Down Syndrome |  |  |  | Down Syndrome |  |  |  |
| --- | --- | --- | --- | --- | --- | --- | --- | --- |
|  | No HSCR |  | HSCR |  | No HSCR |  | HSCR |  |
|  | N | % | N | % | N | % | N | % |
| **Age 0-4** | 9,424,933 |  | 2477 |  | 10,857 |  | 223 |  |
| 1 | 916,378 | 9.7 | 291 | 11.7 | 2,727 | 25.1 | 13* | 5.8 |
| 2-3 | 203,966 | 2.2 | 919 | 37.1 | 2,478 | 22.8 | 57 | 25.6 |
| 4-5 | 32,228 | 0.3 | 515 | 20.8 | 811 | 7.5 | 67 | 30.0 |
| 6-9 | 16,805 | 0.2 | 385 | 15.5 | 538 | 5.0 | 64 | 28.7 |
| 10+ | 12,967 | 0.1 | 169 | 6.8 | 298 | 2.7 | 22 | 9.9 |
| **Age 5-9** | 6,193,771 |  | 1631 |  | 7073 |  | 137 |  |
| 1 | 1,022,169 | 16.5 | 169 | 10.4 | 1,451 | 20.5 | <10* |  |
| 2-3 | 335,598 | 5.4 | 540 | 33.1 | 2,058 | 29.1 | 23 | 16.8 |
| 4-5 | 46,777 | 0.8 | 341 | 20.9 | 935 | 13.2 | 36 | 26.3 |
| 6-9 | 21,181 | 0.3 | 292 | 17.9 | 667 | 9.4 | 46 | 33.6 |
| 10+ | 17,317 | 0.3 | 202 | 12.4 | 406 | 5.7 | 25 | 18.2 |
| **Age 10-14** | 2,907,234 |  | 791 |  | 3334 |  | 64 |  |
| 1 | 535,794 | 18.4 | 86 | 10.9 | 514 | 15.4 |  |  |
| 2-3 | 225,004 | 7.7 | 243 | 30.7 | 923 | 27.7 | <10* |  |
| 4-5 | 36,282 | 1.2 | 157 | 19.8 | 519 | 15.6 | 14 | 21.9 |
| 6-9 | 16,080 | 0.6 | 149 | 18.8 | 454 | 13.6 | 24 | 37.5 |
| 10+ | 12,840 | 0.4 | 110 | 13.9 | 284 | 8.5 | 17 | 26.6 |

\*Grouped with preceding rows and/or number of children with 0 procedures, due to small cell sizes

**Supplementary Table 5:** Most frequently recorded OPCS codes in children with Hirschsprung (for exclusions see Supplementary Table 1).

| OPCS code | Description | N (%) children with code |
| --- | --- | --- |
| **Age 0-4 years, N=2700** |  |  |
| H41.2 | Peranal excision of lesion of rectum | 1989 (73.7) |
| H41.8 | Other operations on rectum through anus (Other, specified) | 1649 (61.1) |
| H19.1 | Open biopsy of lesion of colon | 529 (19.6) |
| G75.3 | Closure of ileostomy | 516 (19.1) |
| H56.8 | Other operations on anus (Other, specified) | 449 (16.6) |
| H44.4 | Examination of rectum under anaesthetic | 389 (14.4) |
| G74.2 | Creation of temporary ileostomy | 386 (14.3) |
| H46.8 | Other operations on rectum (Other, specified) | 323 (12.0) |
| H44.3 | Manual evacuation of impacted faeces from rectum | 320 (11.9) |
| H15.4 | Closure of colostomy | 281 (10.4) |
| **Age 5-9 years, N=1768** |  |  |
| H56.8 | Other operations on anus (Other, specified) | 112 (6.3) |
| H44.3 | Manual evacuation of impacted faeces from rectum | 104 (5.9) |
| H41.2 | Peranal excision of lesion of rectum | 84 (4.8) |
| H44.4 | Examination of rectum under anaesthetic | 80 (4.5) |
| F10.4 | Extraction of multiple teeth | 75 (4.2) |
| G45.1 | Fibreoptic endoscopic examination of upper gastrointestinal tract and biopsy of lesion of upper gastrointestinal tract | 72 (4.1) |
| H22.1 | Diagnostic fibreoptic endoscopic examination of colon and biopsy of lesion of colon | 66 (3.7) |
| D15.1 | Myringotomy with insertion of ventilation tube through tympanic membrane | 47 (2.7) |
| H41.8 | Other operations on rectum through anus, Other specified | 40 (2.3) |
| G75.3 | Closure of ileostomy | 37 (2.1) |
| **Age 10-14 years, N=855** |  |  |
| G45.1 | Fibreoptic endoscopic examination of upper gastrointestinal tract and biopsy of lesion of upper gastrointestinal tract | 31 (3.6) |
| H22.1 | Diagnostic fibreoptic endoscopic examination of colon and biopsy of lesion of colon | 28 (3.3) |
| F10.4 | Extraction of multiple teeth | 26 (3) |
| H44.3 | Manual evacuation of impacted faeces from rectum | 24 (2.8) |
| T41.3 | Freeing of adhesions of peritoneum NEC | 20 (2.3) |
| H41.2 | Peranal excision of lesion of rectum | 19 (2.2) |
| H46.3 | Intubation of rectum for pressure manometry | 18 (2.1) |
| H56.8 | Other operations on anus (Other, specified) | 17 (2) |
| H44.4 | Examination of rectum under anaesthetic | 16 (1.9) |
| G75.3 | Closure of ileostomy | 11 (1.3) |

NEC = Not Elsewhere Classified; NOC = Not Otherwise Classified

**Supplementary Table 6:**
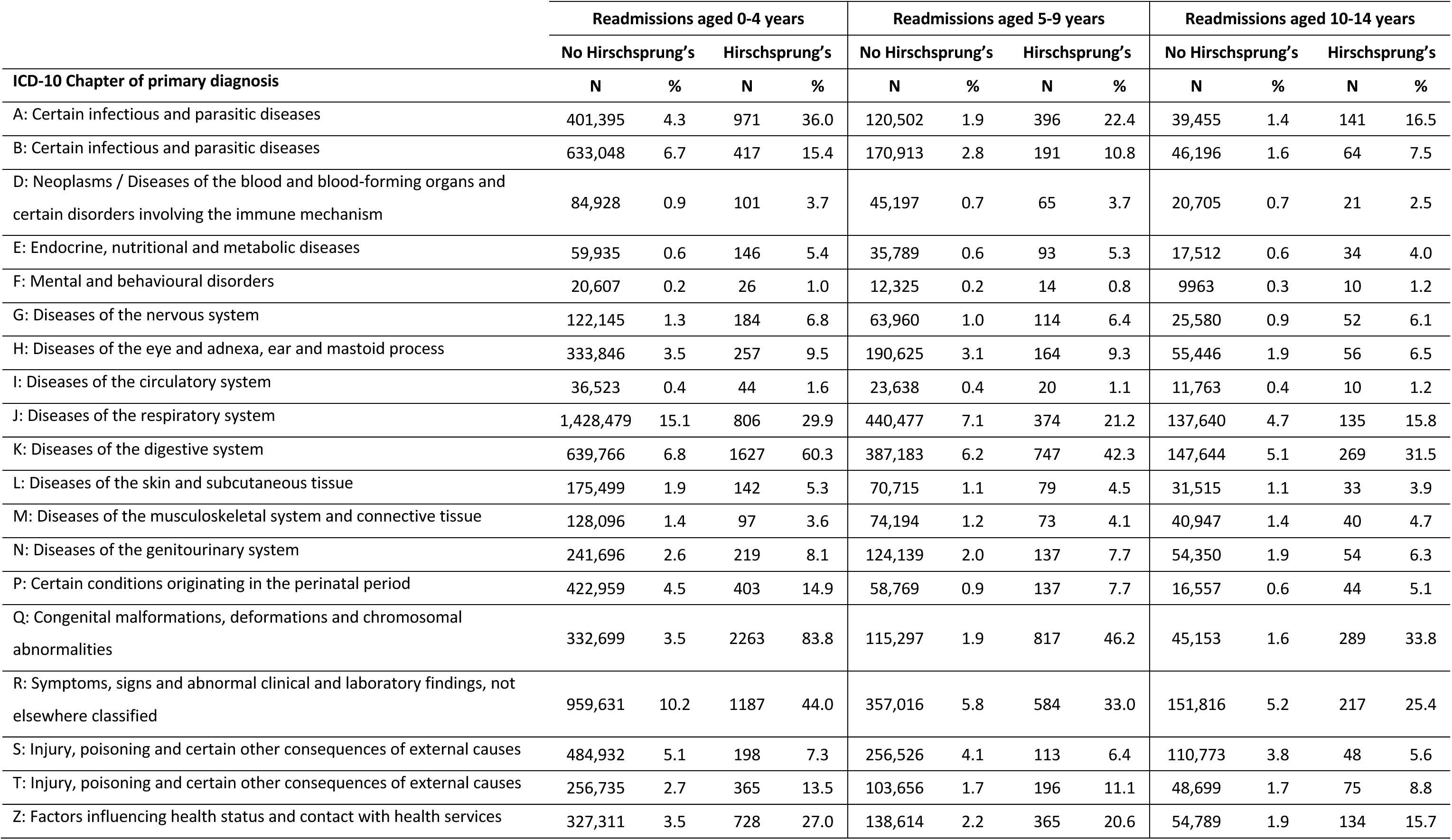
Number and percentage of children admitted age 0-14 years with primary diagnoses within each ICD 10 chapter.

**Supplementary Table 7:** Number and percentage of children admitted age 0-4 years with primary diagnoses within each ICD 10 chapter, according to sex.

|  | Male |  |  |  | Female |  |  |  |
| --- | --- | --- | --- | --- | --- | --- | --- | --- |
|  | No Hirschsprung |  | Hirschsprung |  | No Hirschsprung |  | Hirschsprung |  |
| ICD-10 Chapter of primary diagnosis | N | % | N | % | N | % | N | % |
| A: Certain infectious and parasitic diseases | 215,157 | 4.5 | 739 | 37.6 | 186,238 | 4.1 | 232 | 31.7 |
| B: Certain infectious and parasitic diseases | 373,379 | 7.7 | 304 | 15.5 | 259,669 | 5.7 | 113 | 5.7 |
| D: Diseases of the blood and blood-forming organs and certain disorders involving the immune mechanism | 44,090 | 0.9 | 77 | 3.9 | 40,838 | 0.9 | 24 | 1.2 |
| E: Endocrine, nutritional and metabolic diseases | 31,631 | 0.7 | 103 | 5.2 | 28,304 | 0.6 | 43 | 2.2 |
| G: Diseases of the nervous system | 71,404 | 1.5 | 124 | 6.3 | 50,741 | 1.1 | 60 | 3.1 |
| H: Diseases of the eye and adnexa, ear and mastoid process | 191,402 | 4.0 | 182 | 9.3 | 142,444 | 3.1 | 75 | 3.8 |
| I: Diseases of the circulatory system | 21,183 | 0.4 | 31 | 1.6 | 15,340 | 0.3 | 13 | 0.7 |
| J: Diseases of the respiratory system | 821,919 | 17.0 | 586 | 29.8 | 606,560 | 13.2 | 220 | 11.2 |
| K: Diseases of the digestive system | 363,591 | 7.5 | 1219 | 62.0 | 276,175 | 6.0 | 408 | 20.8 |
| L: Diseases of the skin and subcutaneous tissue | 97,333 | 2.0 | 101 | 5.1 | 78,166 | 1.7 | 41 | 2.1 |
| M: Diseases of the musculoskeletal system and connective tissue | 72,175 | 1.5 | 65 | 3.3 | 55,921 | 1.2 | 32 | 1.6 |
| N: Diseases of the genitourinary system | 153,274 | 3.2 | 154 | 7.8 | 88,422 | 1.9 | 65 | 3.3 |
| P: Certain conditions originating in the perinatal period | 237,626 | 4.9 | 301 | 15.3 | 185,333 | 4.0 | 102 | 5.2 |
| Q: Congenital malformations, deformations and chromosomal abnormalities | 225,384 | 4.7 | 1699 | 86.4 | 107,315 | 2.3 | 564 | 28.7 |
| R: Symptoms, signs and abnormal clinical and laboratory findings, not elsewhere classified | 536,978 | 11.1 | 875 | 44.5 | 422,653 | 9.2 | 312 | 15.9 |
| S: Injury, poisoning and certain other consequences of external causes | 284,480 | 5.9 | 158 | 8.0 | 200,452 | 4.4 | 40 | 2.0 |
| T: Injury, poisoning and certain other consequences of external causes | 143,883 | 3.0 | 267 | 13.6 | 112,852 | 2.5 | 98 | 5.0 |
| Z: Factors influencing health status and contact with health services | 183,979 | 3.8 | 535 | 27.2 | 143,332 | 3.1 | 193 | 9.8 |

**Supplementary Table 8:** Percentage of children readmitted age 5-9 years with primary diagnoses within each ICD 10 chapter, according to sex.

| ICD-10 Chapter of primary diagnosis | Male |  |  |  | Female |  |  |  |
| --- | --- | --- | --- | --- | --- | --- | --- | --- |
|  | No Hirschsprung |  | Hirschsprung |  | No Hirschsprung |  | Hirschsprung |  |
|  | N | % | N | % | N | % | N | % |
| A: Certain infectious and parasitic diseases | 66,632 | 2.1 | 313 | 24.1 | 53,870 | 1.8 | 83 | 17.7 |
| B: Certain infectious and parasitic diseases | 101,933 | 3.2 | 137 | 10.5 | 68,980 | 2.3 | 54 | 11.5 |
| E: Endocrine, nutritional and metabolic diseases | 23,799 | 0.8 | 50 | 3.8 | 21,398 | 0.7 | 15 | 3.2 |
| G: Diseases of the nervous system | 36,656 | 1.2 | 79 | 6.1 | 27,304 | 0.9 | 35 | 7.5 |
| H: Diseases of the eye and adnexa, ear and mastoid process | 104,649 | 3.3 | 118 | 9.1 | 85,976 | 2.8 | 46 | 9.8 |
| J: Diseases of the respiratory system | 252,979 | 8.0 | 284 | 21.9 | 187,498 | 6.2 | 90 | 19.2 |
| K: Diseases of the digestive system | 213,906 | 6.7 | 568 | 43.7 | 173,277 | 5.7 | 179 | 38.2 |
| M: Diseases of the musculoskeletal system and connective tissue | 39,604 | 1.2 | 60 | 4.6 | 31,111 | 1.0 | 19 | 4.1 |
| N: Diseases of the genitourinary system | 41,385 | 1.3 | 51 | 3.9 | 32,809 | 1.1 | 22 | 4.7 |
| Q: Congenital malformations, deformations and chromosomal abnormalities | 85,809 | 2.7 | 98 | 7.5 | 38,330 | 1.3 | 39 | 8.3 |
| R: Symptoms, signs and abnormal clinical and laboratory findings, not elsewhere classified | 35,488 | 1.1 | 108 | 8.3 | 23,281 | 0.8 | 29 | 6.2 |
| S: Injury, poisoning and certain other consequences of external causes | 78,147 | 2.5 | 621 | 47.8 | 37,150 | 1.2 | 196 | 41.8 |
| T: Injury, poisoning and certain other consequences of external causes | 202,774 | 6.4 | 439 | 33.8 | 154,242 | 5.1 | 145 | 30.9 |
| Z: Factors influencing health status and contact with health services | 152,010 | 4.8 | 90 | 6.9 | 104,516 | 3.5 | 23 | 4.9 |

**Supplementary Table 9:** Number and percentage of children readmitted for specific diagnoses, according to the most frequently occurring ICD10 codes in primary diagnosis fields, stratified by sex.

|  |  | No Hirschsprung |  |  |  | Hirschsprung |  |  |  |
| --- | --- | --- | --- | --- | --- | --- | --- | --- | --- |
| Primary diagnosis |  | N | % | N | % | N | % | N | % |
| Readmissions 0-4 years |  | Male (N=4,829,738) |  | Female (N=4,593,576) |  | Male (N=1966) |  | Female (N=733) |  |
| K590 | Constipation | 45242 | 0.9 | 42793 | 0.9 | 689 | 35.0 | 243 | 33.2 |
| A099 | Gastroenteritis and colitis of unspecified origin | 48762 | 1.0 | 41988 | 0.9 | 405 | 20.6 | 136 | 18.6 |
| K529 | Noninfective gastroenteritis and colitis, unspecified | 60078 | 1.2 | 50136 | 1.1 | 367 | 18.7 | 111 | 15.1 |
| R11 | Nausea and vomiting | 58201 | 1.2 | 51974 | 1.1 | 322 | 16.4 | 104 | 14.2 |
| A084 | Viral intestinal infection, unspecified | 117574 | 2.4 | 106920 | 2.3 | 309 | 15.7 | 104 | 14.2 |
| B349 | Viral infection, unspecified | 352891 | 7.3 | 239897 | 5.2 | 281 | 14.3 | 100 | 13.6 |
| J069 | Acute upper respiratory infection, unspecified | 259209 | 5.4 | 202818 | 4.4 | 248 | 12.6 | 99 | 13.5 |
| R14 | Flatulence and related conditions | 1807 | 0.0 | 1743 | 0.0 | 199 | 10.1 | 57 | 7.8 |
| K914 | Colostomy and enterostomy malfunction | 880 | 0.0 | 682 | 0.0 | 194 | 9.9 | 62 | 8.5 |
| R104 | Other and unspecified abdominal pain | 49089 | 1.0 | 44756 | 1.0 | 165 | 8.4 | 66 | 9.0 |
| J22 | Unspecified acute lower respiratory infection | 126195 | 2.6 | 102445 | 2.2 | 150 | 7.6 | 67 | 9.1 |
| Readmissions 5-9 years |  | Male (N=3,170,930) |  | Female (N=3,018,246) |  | Male (N=1299) |  | Female (N=469) |  |
| K590 | Constipation | 33617 | 1.1 | 31825 | 1.1 | 474 | 36.5 | 163 | 34.8 |
| K529 | Noninfective gastroenteritis and colitis, unspecified | 59145 | 1.9 | 49460 | 1.6 | 357 | 27.5 | 103 | 22.0 |
| R11 | Nausea and vomiting | 41829 | 1.3 | 37276 | 1.2 | 223 | 17.2 | 66 | 14.1 |
| A084 | Viral intestinal infection, unspecified | 83499 | 2.6 | 74851 | 2.5 | 212 | 16.3 | 61 | 13.0 |
| B349 | Viral infection, unspecified | 207608 | 6.5 | 144628 | 4.8 | 169 | 13.0 | 70 | 14.9 |
| A099 | Gastroenteritis and colitis of unspecified origin | 21452 | 0.7 | 18177 | 0.6 | 165 | 12.7 | 57 | 12.2 |
| J069 | Acute upper respiratory infection, unspecified | 170971 | 5.4 | 131781 | 4.4 | 157 | 12.1 | 62 | 13.2 |
| K914 | Colostomy and enterostomy malfunction | 636 | 0.0 | 474 | 0.0 | 136 | 10.5 | 39 | 8.3 |
| R104 | Other and unspecified abdominal pain | 41955 | 1.3 | 38944 | 1.3 | 129 | 9.9 | 42 | 9.0 |
| R14 | Flatulence and related conditions | 1239 | 0.0 | 1181 | 0.0 | 123 | 9.5 | 42 | 9.0 |
| K590 | Constipation | 33617 | 1.1 | 31825 | 1.1 | 474 | 36.5 | 163 | 34.8 |

| *Readmissions 10-14 years* |  | **Male (N=1,487,034)** |  | **Female (N=1,418,725)** |  | **Male (N=630)** |  | **Female (N=225)** |  |
| --- | --- | --- | --- | --- | --- | --- | --- | --- | --- |
| K590 | Constipation | 16756 | 1.1 | 16041 | 1.1 | 228 | 36.2 | 83 | 36.9 |
| K529 | Noninfective gastroenteritis and colitis, unspecified | 32430 | 2.2 | 26936 | 1.9 | 184 | 29.2 | 51 | 22.7 |
| R11 | Nausea and vomiting | 20492 | 1.4 | 18270 | 1.3 | 101 | 16.0 | 26 | 11.6 |
| A084 | Viral intestinal infection, unspecified | 36000 | 2.4 | 32208 | 2.3 | 94 | 14.9 | 25 | 11.1 |
| J069 | Acute upper respiratory infection, unspecified | 80556 | 5.4 | 61468 | 4.3 | 76 | 12.1 | 26 | 11.6 |
| B349 | Viral infection, unspecified | 73685 | 5.0 | 54935 | 3.9 | 71 | 11.3 | 28 | 12.4 |
| R14 | Flatulence and related conditions | 590 | 0.0 | 557 | 0.0 | 69 | 11.0 | 20 | 8.9 |
| R104 | Other and unspecified abdominal pain | 23976 | 1.6 | 23627 | 1.7 | 62 | 9.8 | 17 | 7.6 |
| K914 | Colostomy and enterostomy malfunction | 288 | 0.0 | 227 | 0.0 | 57 | 9.0 | 20 | 8.9 |
| A09 | Other gastroenteritis and colitis of infectious and unspecified origin | 5610 | 0.4 | 4925 | 0.3 | 54 | 8.6 | 10 | 3.6 |
| Z433 | Attention to colostomy | 254 | 0.0 | 201 | 0.0 | 36 | 5.7 | 19 | 8.4 |

## References

1. Montalva L, et al. Hirschsprung disease. Nature Reviews Disease Primers. 2023;9(1):54.

2. Butler Tjaden NE, Trainor PA. The developmental etiology and pathogenesis of Hirschsprung disease. Transl Res. 2013;162(1):1–15.

3. Ostertag-Hill CA, et al. Late Diagnosis of Hirschsprung Disease: Clinical Presentation and Long-Term Functional Outcomes. Journal of pediatric surgery. 2024;59(2):220–4.

4. Langer JC. Surgical approach to Hirschsprung disease. Semin Pediatr Surg. 2022;31(2):151156.

5. Jensen AR, Frischer JS. Surgical history of Hirschsprung disease. Semin Pediatr Surg. 2022;31(2):151174.

6. Davidson JR, et al. Comparative cohort study of Duhamel and endorectal pull-through for Hirschsprung’s disease. BJS Open. 2022;6(1).

7. Davidson JR, et al. Outcomes in Hirschsprung’s disease with coexisting learning disability. Eur J Pediatr. 2021;180(12):3499–507.

8. Damkjær M, et al. Children with Hirschsprung’s disease have high morbidity in the first 5 years of life. Birth Defects Research. 2024;116(5):e2338.

9. Lu C, et al. Bowel function at preschool and early childhood age in children with long-segment Hirschsprung disease. European Journal of Pediatrics. 2023;182(3):1251–9.

10. Bradnock TJ, et al. Hirschsprung’s disease in the UK and Ireland: incidence and anomalies. Arch Dis Child. 2017;102(8):722–7.

11. Davidson JR, et al. Long-term surgical and patient-reported outcomes of Hirschsprung Disease. J Pediatr Surg. 2021;56(9):1502–11.

12. Allin BSR, et al. NETS(1HD) study: development of a Hirschsprung’s disease core outcome set. Arch Dis Child. 2017;102(12):1143–51.

13. Jarvi K, et al. Bowel Function and Gastrointestinal Quality of Life Among Adults Operated for Hirschsprung Disease During Childhood: A Population-Based Study. Annals of surgery. 2010;252(6).

14. Shah R. Anesthesia exposure in the young child and long-term cognition: an integrated review. AANA journal. 2019;87(3):231–41.

15. Roorda D, et al. Neurodevelopmental outcome of patients with congenital gastrointestinal malformations: a systematic review and meta-analysis. Archives of Disease in Childhood-Fetal and Neonatal Edition. 2021;106(6):635–42.

16. Glatz P, et al. Association of Anesthesia and Surgery During Childhood With Long-term Academic Performance. JAMA pediatrics. 2017;171(1):e163470-e.

17. McCann ME, et al. Neurodevelopmental outcome at 5 years of age after general anaesthesia or awake-regional anaesthesia in infancy (GAS): an international, multicentre, randomised, controlled equivalence trial. Lancet. 2019;393(10172):664–77.

18. Mc Grath-Lone L, et al. Data Resource Profile: The Education and Child Health Insights from Linked Data (ECHILD) Database. Int J Epidemiol. 2021;dyab149, 10.1093/ije/dyab149.

19. Jay M, et al. Data Resource Profile: the National Pupil Database. Int J Popul Data Sci. 2019;4(1).

20. Herbert A, et al. Data Resource Profile: Hospital Episode Statistics Admitted Patient Care (HES APC). Int J Epidemiol. 2017;Epub:doi: 10.1093/ije/dyx015.

21. Feng Q, et al. Data Resource Profile: a national linked mother-baby cohort of health, education and social care data in England (ECHILD-MB). Int J Epidemiol. 2024;53(3).

22. Nasr A, et al. Validation of algorithms to determine incidence of Hirschsprung disease in Ontario, Canada: a population-based study using health administrative data. Clinical Epidemiology. 2017;9(null):579–90.

23. Giuliani S, et al. Outcomes of primary versus multiple-staged repair in hirschsprung’s disease in england. European Journal of Pediatric Surgery. 2020;30(01):104–10.

24. Jay MA, Gilbert R. Special educational needs, social care and health. Arch Dis Child. 2021;106(1):83–5.

25. Cole TJ, et al. Birth weight and longitudinal growth in infants born below 32 weeks’ gestation: a UK population study. Arch Dis Child Fetal Neonatal Ed. 2014;99(1):F34–F40.

26. Paus T, et al. Why do many psychiatric disorders emerge during adolescence? Nature Reviews Neuroscience. 2008;9(12):947–57.

27. Scheiner C, et al. Mental disorders at the beginning of adolescence: Prevalence estimates in a sample aged 11-14 years. Public Health Pract (Oxf). 2022;4:100348.

28. Allin BSR, et al. Outcomes at five to eight years of age for children with Hirschsprung’s disease. Arch Dis Child. 2021;106(5):484–90.

29. Meinds RJ, et al. Long-term functional outcomes and quality of life in patients with Hirschsprung’s disease. Br J Surg. 2019;106(4):499–507.

30. Pakarinen MP, Mutanen A. Long-term outcomes and quality of life in patients with Hirschsprung disease. World J Pediatr Surg. 2024;7(3):e000859.

31. Dai Y, et al. Long-term outcomes and quality of life of patients with Hirschsprung disease: a systematic review and meta-analysis. BMC Gastroenterol. 2020;20(1):67.

32. Thompson DS, et al. Transitional Care in Patients With Hirschsprung Disease: Those Left Behind. Dis Colon Rectum. 2024;67(7):977–84.

33. Allin BSR, et al. Impact of rectal dissection technique on primary-school-age outcomes for a British and Irish cohort of children with Hirschsprung disease. J Pediatr Surg. 2022;57(12):902–11.

34. Balela N, et al. Prognostic factors for persistent obstructive symptoms in patients with Hirschsprung disease following pull-through. PloS one. 2023;18(9):e0290430.

35. Hou J, et al. Robotic-assisted Swenson procedure for Hirschsprung’s disease with a median age of 35 days: a single-center retrospective study. Pediatr Surg Int. 2025;41(1):87.

36. Li Y, et al. Comparison of robot-assisted and laparoscopic-assisted modified Soave short muscle cuff anastomosis surgeries for classical Hirschsprung disease. BMC Surg. 2025;25(1):78.

37. Karlsen RA, et al. Comparison of clinical outcomes after total transanal and laparoscopic assisted endorectal pull-through in patients with rectosigmoid Hirschsprung disease. J Pediatr Surg. 2022;57(9):69–74.

38. Gely Y, et al. Surgical Outcomes for Patients With Trisomy 21 and Hirschsprung’s Disease: An NSQIP-Pediatric Study. J Surg Res. 2024;302:724–31.

39. Kyrklund K, et al. ERNICA guidelines for the management of rectosigmoid Hirschsprung’s disease. Orphanet J Rare Dis. 2020;15(1):164.

40. Rintala RJ, Pakarinen MP. Long-term outcomes of Hirschsprung’s disease. Semin Pediatr Surg. 2012;21(4):336–43.

41. Nasr A, et al. Long-term Outcomes of Patients Surgically Treated for Hirschsprung Disease. J Can Assoc Gastroenterol. 2021;4(5):201–6.

42. Collins L, et al. Quality of life outcomes in children with Hirschsprung disease. J Pediatr Surg. 2017;52(12):2006–10.

43. Svetanoff WJ, et al. Psychosocial factors affecting quality of life in patients with anorectal malformation and Hirschsprung disease-a qualitative systematic review. J Pediatr Surg. 2022;57(3):387–93.

44. Verkuijl SJ, et al. Familial Experience With Hirschsprung’s Disease Improves the Patient’s Ability to Cope. Front Pediatr. 2022;10:820976.

45. Huang HB, et al. A systematic review of school functioning in pediatric patients with Hirschsprung disease. Pediatr Surg Int. 2025;41(1):123.

46. Montalva L, et al. Hirschsprung disease. Nat Rev Dis Primers. 2023;9(1):54.

47. Moore SW. Advances in understanding the association between Down syndrome and Hirschsprung disease (DS-HSCR). Pediatr Surg Int. 2018;34(11):1127–37.

48. Elgendy MM, et al. Prevalence and Outcomes of Gastrointestinal Anomalies in Down Syndrome. Am J Perinatol. 2024;41(15):2047–52.

49. Granström AL, et al. Population-based study shows that Hirschsprung disease does not have a negative impact on education and income. Acta Paediatr. 2016;105(12):1508–12.

50. de Beaufort CMC, et al. Transitional Care for Patients with Congenital Colorectal Diseases: An EUPSA Network Office, ERNICA, and eUROGEN Joint Venture. J Pediatr Surg. 2023;58(12):2319–26.

51. Plascevic J, et al. Transitional Care in Anorectal Malformation and Hirschsprung’s Disease: A Systematic Review of Challenges and Solutions. J Pediatr Surg. 2024;59(6):1019–27.

